# Assessment of the impact of the Vaccine Pass Policy on COVID-19 vaccine hesitancy and uptake among Chinese adults in Hong Kong

**DOI:** 10.1101/2023.12.03.23299354

**Authors:** Irene O. L. Wong, Cherry Wong, Nelly Mak, Alan Dai, Jingyi Xiao, Peng Wu, Michael Y. Ni, Qiuyan Liao, Benjamin J. Cowling

## Abstract

**Background:** Recognising the importance of attaining high vaccine coverage to mitigate the COVID-19 impact, a Vaccine Pass scheme was implemented during and after the first large Omicron wave in Hong Kong in early 2022 requiring three doses by June 2022. We evaluated the impact of the Vaccine Pass policy on vaccine uptake in adults.

**Methods:** We analyzed patterns in vaccine uptake and hesitancy using local data from the population vaccine registry and 32 cross-sectional surveys conducted from October 2021 to December 2022. We examined the association of Vaccine Pass phases with vaccine uptake, taking into account covariables including self-risk perception, perceived self-efficacy in preventing COVID-19 and trust in government in pandemic control as well as physical distancing measures and demographics.

**Findings:** The uptake of primary series and third doses was significantly associated with stages of Vaccine Pass implementation, and other statistically significant drivers included age group, chronic condition, higher perceived personal susceptibility to COVID-19, higher trust in government, and higher educational attainment. Older adults (≥65y) were less likely to be vaccinated against COVID-19, compared to adults aged 18-44 years.

**Interpretation:** Vaccine uptake in older adults was observed to have increased by a greater extent after the policy announcement and implementation, which occurred during and after a large Omicron wave with high mortality in older ages in early 2022. Since the policy withdrawal the uptake of further booster doses has been very low in all ages. Improving voluntary booster uptake in older adults should be prioritized.

**Funding:** Health and Medical Research Fund, Hong Kong.

## Introduction

The roll-out of COVID-19 vaccines around the world in 2021 allowed safer relaxation of public health and social measures that had caused enormous social and economic disruption in many parts of the world. In many higher income locations, serologic studies indicated that the cumulative incidence of infections remained low until after vaccination programs had started (1). Locations which more quickly achieved higher vaccine uptake had lower subsequent COVID-19 mortality (2). In Hong Kong, two COVID-19 vaccines were available from February 2021, the mRNA vaccine BNT162b2 (BioNTech/Fosun Pharma/Pfizer) and the inactivated vaccine CoronaVac (Sinovac). Individuals were able to choose which vaccine they received for their primary series and could choose the same or different vaccine for booster doses. Initial prioritization for vaccines was for older adults and healthcare workers, with progressive broadening of availability to other age groups in early 2021. All vaccinations were provided for free through vaccination centres established across the territory.

Following moderate uptake of vaccinations initially, at a time when COVID-19 was being contained effectively by public health measures, a number of schemes were introduced by the government and by private organizations to encourage vaccination. During this period, arriving travelers were required to undergo quarantine in designated hotels for 14 or 21 days (3). From 12 May to 19 August 2021, vaccinated individuals were permitted a 7-day reduction in this period (3). Various organizations created lotteries in which only vaccinated individuals could participate (4). On 2 August 2021, following repeated and prolonged periods of school closures and online education during the pandemic (5), the government announced that whole-day schooling could resume in any schools that achieved at least 70% two-dose uptake among staff and students (4). Some restrictions were also introduced, including for example a requirement that unvaccinated individuals undergo asymptomatic COVID-19 testing more frequently and pay for their own tests (4).

While control measures were successful in containing COVID-19 for two years (5), the spread of the Omicron variant could not be contained in early 2022 resulting in high levels of hospitalization and mortality in March-April 2022, with almost all severe and fatal cases occurring in unvaccinated older adults (6). Recognising that achieving high vaccine coverage was important to mitigate the impact of COVID-19, particularly during and after the impact of this large Omicron wave, a Vaccine Pass scheme was announced on 9 February 2022 and implemented on 24 February 2022 (7). Under this scheme, similar to vaccine passports applied in other parts of the world (8), receipt of a specified number of vaccine doses was a condition for adults to access various locations including restaurants, fitness centres, shopping malls, government offices, schools, universities, and libraries. For individuals working in these locations, compliance with the Vaccine Pass criteria became a condition of continued employment and anti-discrimination legislation was modified to allow COVID-19 vaccination status to be a valid reason for dismissal. The Vaccine Pass scheme was progressively broadened through 2022, including a requirement for a third dose by June 2022, until the scheme was eventually discontinued on 29 December 2022.

Here, we analyse patterns in vaccine uptake and vaccine hesitancy in adults in Hong Kong based on analysis of the local population vaccine registry along with data from 32 telephone interview surveys from October 2021 through to December 2022. Our objective is to explore the potential impact of Vaccine Pass policies on vaccine uptake in adults of different ages.

## Methods

### Sources of data

We retrieved population denominators in 2021 from the Census and Statistics Department. We also obtained local population-based data on age-specific vaccination status from the Centre for Health Protection of the Department of Health on weekly basis from late 2021 to 2022. All COVID-19 vaccine doses administered in Hong Kong were recorded in this registry, and residents were also able to register any doses received overseas. We also collected official data on the number of daily laboratory-confirmed COVID-19 cases and COVID-19 related deaths in Hong Kong during our study period.

We analyzed data from a series of community-based telephone interview surveys in Hong Kong from October 2021 to December 2022 (9). Serial population-based cross-sectional surveys were conducted using random-digit dialling of both land-based and mobile telephone numbers approximately in the ratio of 1:1. Survey data were recorded by a Web-based Computer Assisted Telephone Interviewing system. The target population included Cantonese-speaking Hong Kong residents (excluding travelers to Hong Kong) aged ≥18 years of age who have landline or mobile telephone numbers. In the present study, we used data from a total of 32 survey rounds, conducted every 2-4 weeks each with sample sizes of either approximately 500 or 1000 respondents. Our study protocol was approved by the Institutional Research Board of the University of Hong Kong (ref: US 20-095).

### Outcome measures

Our primary outcomes were *COVID-19 vaccination hesitancy* and *self-reported vaccine uptake*. In the telephone surveys, we obtained self-reported COVID-19 vaccination status from the respondents. We also assessed intention to receive a COVID-19 vaccine dose among the unvaccinated respondents from June 2020 through to early July 2022. Receipt of second doses closely tracked receipt of the first dose (10) and was not explored separately. Similarly, we also assessed intention to receive a third/booster COVID-19 vaccine dose for fully vaccinated respondents (i.e., received 2 doses) from in the phase 3 of the Vaccine Pass period from mid-July 2022 to December 2022. The response scale was 7-point categories ranging from “certain”, “very likely”, “likely”, “evens”, “unlikely”, “very unlikely” or “never”. Respondents were defined as having vaccine hesitancy if their likelihood to receive vaccination was “never”, “very unlikely”, “unlikely” or “evens” (11). The primary outcome measure was vaccine hesitancy defined as never, very unlikely, unlikely, or evens to get vaccinated, rather than certain, very likely or likely to receive vaccination during the survey period (11).

Other measures relevant to the present study were also assessed in the survey with a length of 10-15 minutes. For *risk perception of COVID-19,* respondents were asked about their perception of the severity of COVID-19 for themselves if they were infected on a five-point categorical scale ranging from “very mild” to “very serious” (perceived severity), perceived personal likelihood of contracting COVID-19 on a seven-point scale ranging from “never” to “certain” (perceived susceptibility), and worry about contracting COVID-19 on a five-point scale ranging from “not at all worried” to “extremely worried” (worry about contacting COVID-19) in the month following the survey. *Perceived self-efficacy* in preventing COVID-19 was assessed with a statement: “I’m confident that I can take measures to protect myself against novel coronavirus pneumonia.” *Trust in government* in controlling COVID-19 was assessed with a statement: “I trust that the Hong Kong government can take effective measures to control novel coronavirus pneumonia spread in Hong Kong.” (these two response scales ranging from “1=Strongly disagree” to “5= Strongly agree”). We also included a measure of compliance with social gathering avoidance, noting that this measure was previously found to be the best proxy indicator for compliance with physical distancing measures in general (9).

### Statistical analyses

We used joinpoint regression analyses to investigate temporal changes in trends in vaccine uptake of primary series and third doses of COVID-19 vaccinations in Hong Kong each month from October 2021 through to December 2022 by age group based on the vaccine registry data. We quantified the trend in terms of monthly percentage change and average monthly percentage change. We then explored temporal patterns in prevalence of COVID-19 vaccine hesitancy for primary series among all participants from October 2021 to July 2022, and COVID-19 booster vaccine hesitancy from mid July to December 2022. Survey proportions were weighted by applying rim-weighting factors including age, sex, education level and occupation status according to the adult census population distribution in 2019 in Hong Kong, with 95% confidence interval (CI) based on a normal approximation. We used multivariable logistic regression analyses to identify factors associated with self-reported vaccine uptake using the survey data from October 2021 through to July 2022. In the model, we studied the time period effect of various Vaccine Pass phases in 2022 (i.e., pre-annoucement before Febuary 8, post-annoucement on Feburary 8-24, plus full implementation of 3 phases starting from phase 1 on Febuary 24, phase 2 on April 30, phase 3 on May 31 and onwards) in Hong Kong with adjustment for other covariables including self-risk perception measures, perceived control, compliance with physical distancing measure, and demographics. Data analyses were conducted using R 4.2.1 (R Foundation for Statistical Computing, Vienna, Austria) and the Joinpoint Regression Program version 4.2.2 (Statistical Methodology and applications branch, surveillance research program, National Cancer Institute).

## Results

The timeline of the Vaccine Pass scheme is illustrated in **Figure 1** with additional details provided in **Supplementary Table 1**. We analysed data from thirty-two rounds of telephone surveys from 1 October 2021 to 31 December 2022 including a total of 25,012 adult respondents (**Supplementary Figure 1**). Characteristics of survey respondents for the 32 rounds are reported in **Table 1**.

**Figure 1.**
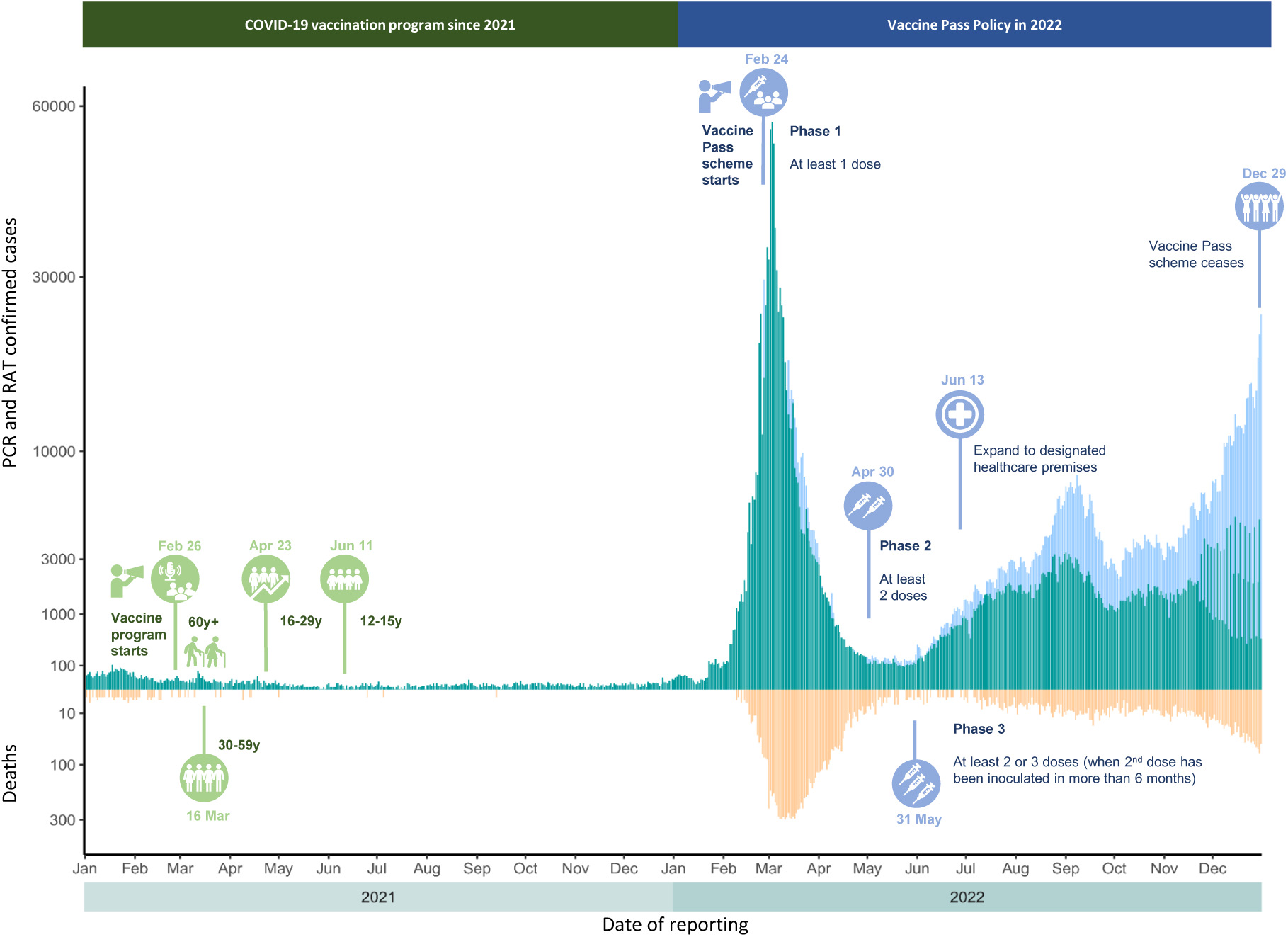
Age coverage of the COVID-19 vaccination program since 2021, Vaccine Pass Policy timeline (From February to December 2022) and local risk (in terms of epidemic curves of PCR-confirmed (green bars) and RAT-confirmed (pastel blue bars) new COVID-19 cases and COVID-related deaths (orange bars) based on official registry data) over time in Hong Kong

**Table 1.** Sample demographic characteristics in the analysis (n=25012)

| Characteristic | Number | % |
| --- | --- | --- |
| Age group |  |  |
| 18-44 years | 7285 | 29.1 |
| 45-64 years | 8043 | 32.2 |
| 65 years or above | 9091 | 36.3 |
| Sex |  |  |
| Male | 9921 | 39.7 |
| Female | 15091 | 60.3 |
| Educational attainment |  |  |
| Primary or lower | 4894 | 19.6 |
| Secondary | 11135 | 44.5 |
| Tertiary or above | 8625 | 34.5 |
| Self-reported doctor-diagnosed chronic condition |  |  |
| No | 15451 | 61.8 |
| Yes | 8851 | 35.4 |

† 593 (2.4%) respondents did not report their age; 358 (1.4%) respondents did not report their education level; 710 (2.8%) respondents did not report whether they had any chronic condition.

Figure 2 shows trajectories over time of 2-dose and 3-dose vaccine uptake rates in Hong Kong by age group. Vaccine uptake for primary series and booster doses continued to increase over time in 2022, and a number of trajectory changes were identified in 2022 in the joinpoint analysis. After the implementation of the Vaccine Pass scheme coinciding with large numbers of COVID-19 deaths in older adults during a large Omicron wave in March-April 2022, there was a faster increase of vaccine uptake in the oldest-old group (≥80 years), compared to other ages. Most of the estimated joinpoints (solid triangles) or the statistically significant change in trends coincided with the dates of the implementation of phase 1 and phase 3 of the Vaccine Pass scheme, with the general pattern being deceleration in uptake after the relevant cut-off date, indicating that the period prior to a cut-off had been associated with faster uptake rates.

**Figure 2.**
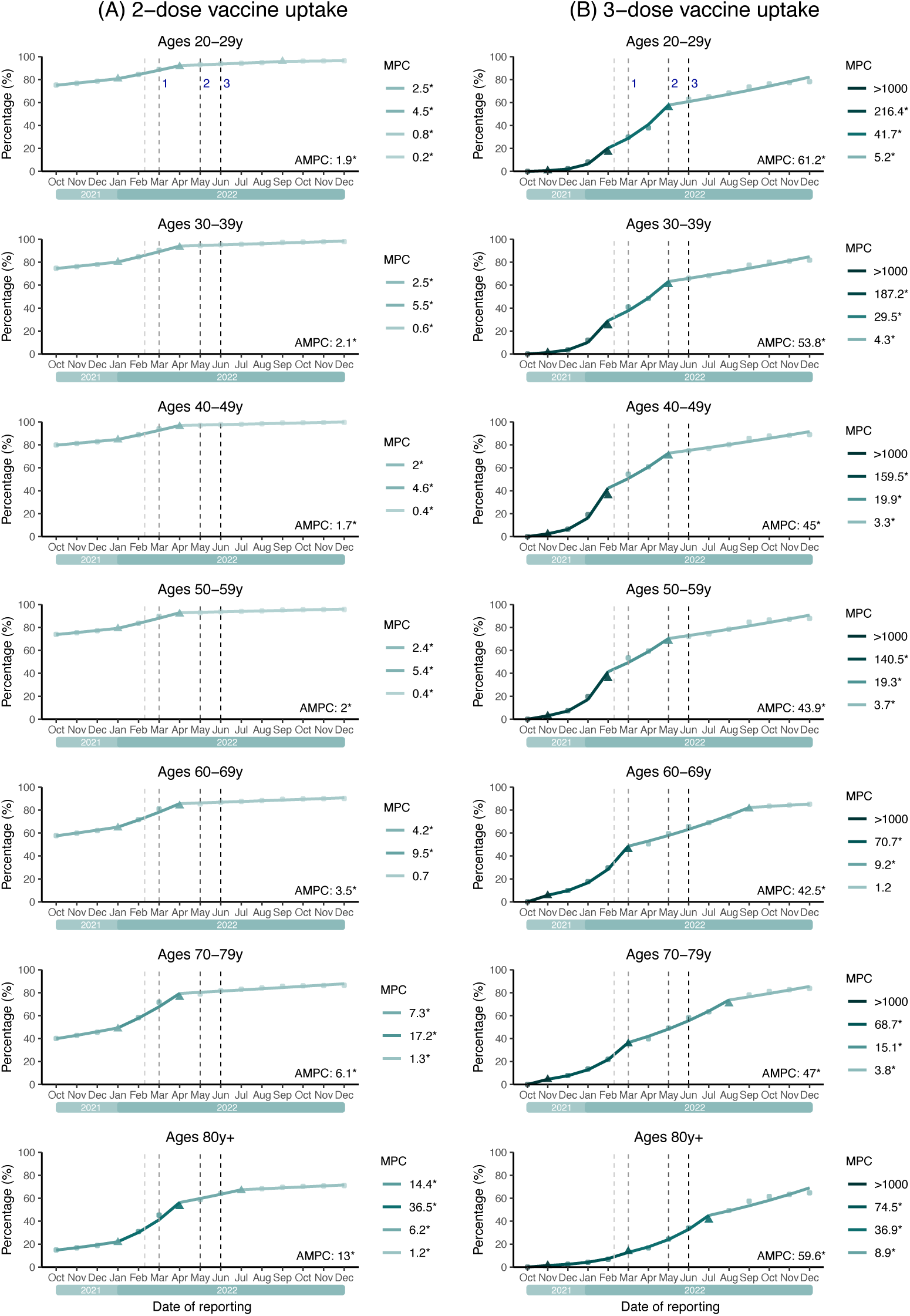
Vaccine uptake in Hong Kong over date of reporting by age group based on official government data with population coverage. The panels A and B show percentages of 2-dose vaccine uptake and percentages of 3-dose vaccine uptake respectively. Notes: 1^st^ vertical dashed line=Vaccine Pass annouces; 2^nd^ vertical dashed line=Phase 1 Vaccine Pass implements with the first dose required; 3^rd^ vertical dashed line=Phase 2 of Vaccine Pass commences with the second dose required; 4^th^ vertical dashed line=Phase 3 of Vaccine Pass commences with the third dose required if ≥6m have elapsed

Figure 3 shows prevalence of vaccine hesitancy for the next dose by age group (18-44, 45-64y, ≥65y) among adult individuals in Hong Kong from October 2021 to July 2022, encompassing the large Omicron epidemic wave in March-April 2022. There was clear decline of vaccine hesitancy in late February 2022 (i.e. just after the Vaccine Pass was implementated) in all three age groups, and hesitancy dropped particularly rapidly in older adults. Figure 4 displays prevalence of booster vaccine hesitancy among fully vaccinated individuals (i.e. at least two doses) in Hong Kong from mid-July to December 2022. Levels of hesitancy were relatively less stagnant after mid-July among the young vaccinated group (from 6.1% in July to 11.1% in December), while the rates did not change substantially after mid-July in older adults who had been fully vaccinated.

**Figure 3.**
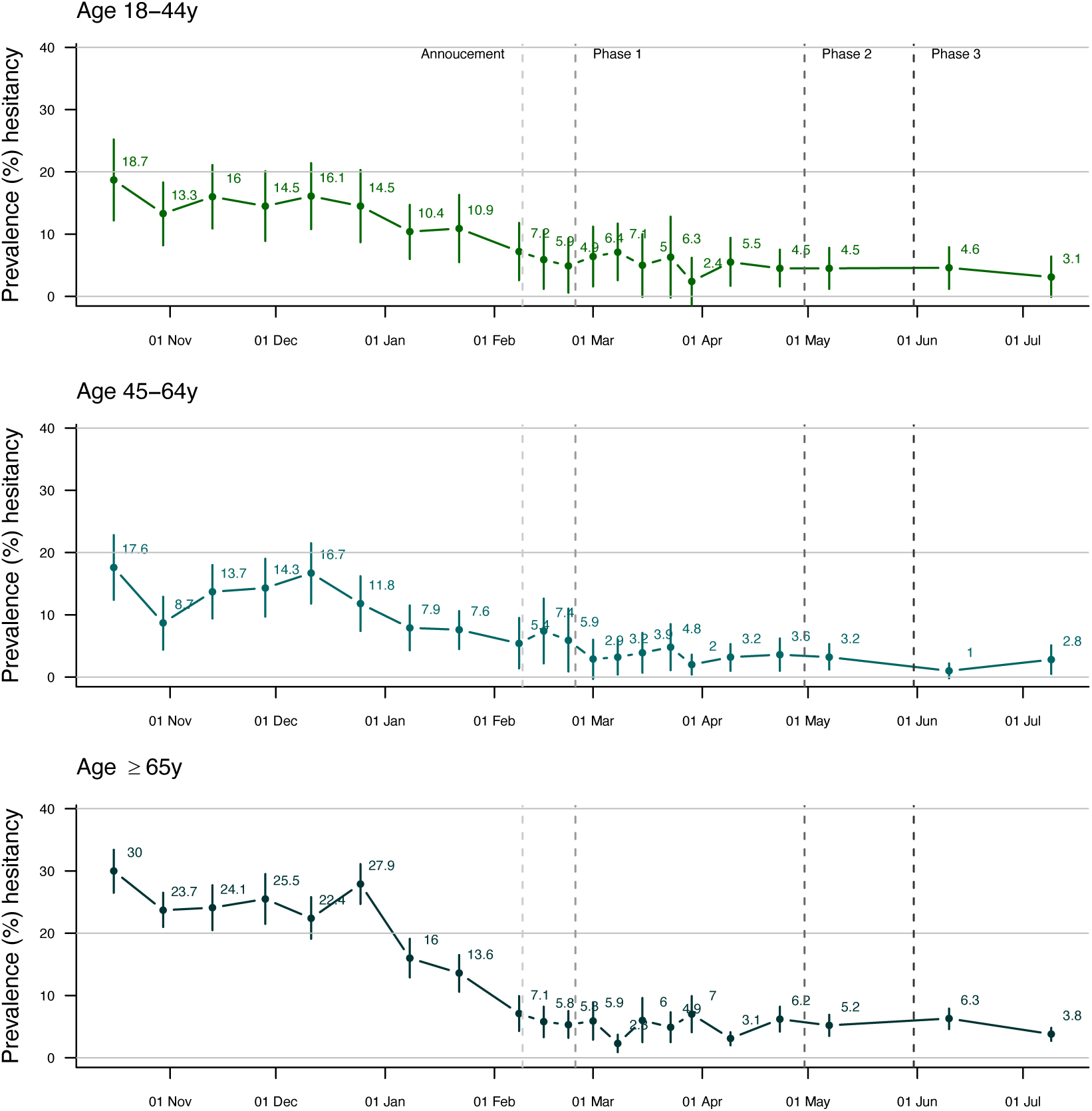
Prevalence of vaccine hesitancy (the first or the next dose) among adult individuals in Hong Kong from October in 2021 to July in 2022 (before and during the 5^th^ wave with omicron main variant circulating), by younger (18-44y, 45-64y) and older (65y+) age group. The prevalence was estimated by the proportion of COVID-19 vaccine hesitancy in each survey, where the proportion was calculated by dividing the number of vaccine hesitants by total number of the respective participants (/overall sample size) in the survey.The proportion was weighted by applying rim-weighting factors including age, sex, education level and occupation status according to the adult census population distribution in 2019 in Hong Kong, with 95% confidence intervals (CIs) by using normal approximation.(Notes: We initially assessed intention to receive a COVID-19 vaccine with 7-point categories ranging from “certain”, “very likely”, “likely”, “evens”, “unlikely”, “very unlikely” or “never”, and we recategoried the scales to define vaccine hesitancy. Respondents were defined as having vaccine hesitancy if their responses would “never” or “very unlikely” or unlikely” / “even” to get vaccinated rather than “very likely” or “likely to vaccinate” during the survey period or vaccinated. Vaccine hesitant was defined as “never”, “very unlikely”, “unlikely” or “evens” to get COVID-19 vaccination, rather than “very likely” or “likely” to vaccinate during the survey period or vaccinated.)

**Figure 4.**
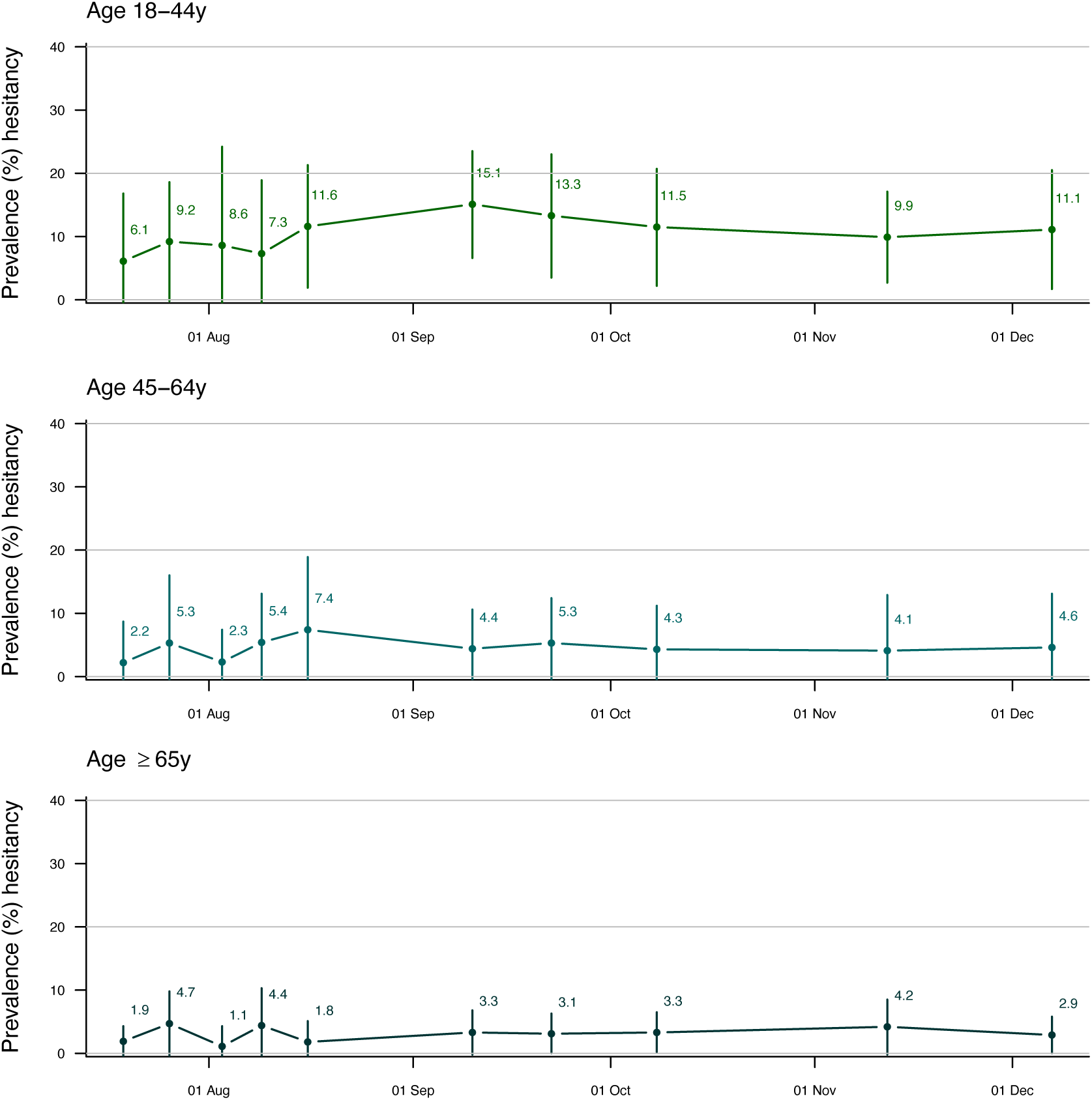
Prevalence of booster vaccine hesitancy (the third dose) among adult individuals in Hong Kong from mid-July to December in 2022 (after the 5^th^ wave with omicron main variant circulating), by younger (18-44y, 45-64y) and older (65y+) age group. The prevalence was estimated by the proportion of COVID-19 vaccine hesitancy in each survey, where the proportion was calculated by dividing the number of vaccine hesitants (2 dose only and hesitant to get the third dose) by total number of the fully vaccinated (self-reported receiving at least two doses) participants in the survey. The proportion was weighted by applying rim-weighting factors including age, sex, education level and occupation status according to the adult census population distribution in 2019 in Hong Kong, with 95% confidence intervals (CIs) by using normal approximation.(Notes: We initially assessed intention to receive a COVID-19 vaccine with 7-point categories ranging from “certain”, “very likely”, “likely”, “evens”, “unlikely”, “very unlikely” or “never”, and we recategoried the scales to define vaccine hesitancy. Respondents were defined as having vaccine hesitancy if their responses would “never” or “very unlikely” or unlikely” / “even” to get vaccinated rather than “very likely” or “likely to vaccinate” during the survey period. Vaccine hesitant was defined as “never”, “very unlikely”, “unlikely” or “evens” to get COVID-19 vaccination, rather than “very likely” or “likely” to vaccinate during the survey period.)

In multivariable regression models using data from October 2021 to July 2022, vaccine uptake was significantly associated with the Vaccine Pass policy in various stages (i.e., post announcement, phase 1 to phase 3) in adults overall, during the pre- and post-omicron period (Figure 5). Compared to a younger age group (18-44y), older adults (≥65y) were statistically significantly less likely to be vacinated against COVID-19. Having a self-reported chronic condition was also associated with a lower chance to receive COVID-19 vaccination. Nevertheless, these two correlated variables, older age and chronic conditions, appeared to be the most crucial factors negatively associated with vacine uptake. On the other hand, higher perceived susceptibility of contracting COVID-19, higher self-perceived external control and higher education were statistically significantly positively associated with vaccine uptake.

**Figure 5.**
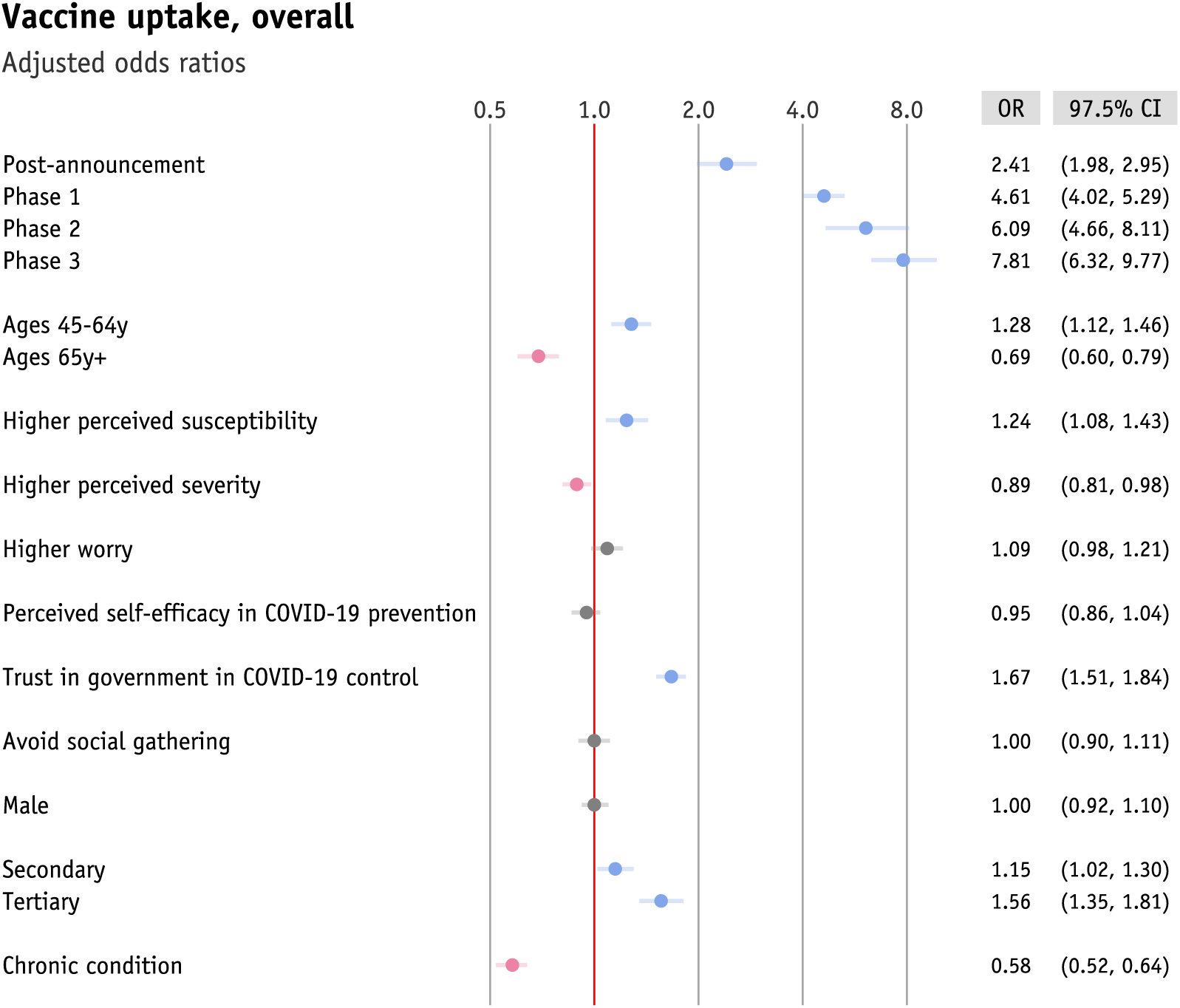
The impact of Vaccine Pass implementation on and factors associated with vaccine uptake (any dose) among adults overall and by various Vaccine Pass phases from pre-announcement to full implementation (from October 2021 to July in 2022 in Hong Kong (during the 5^th^ wave with omircon main variant circulating), by using multivariable logistic regression analysis. The solid circle and horizontal bar as well as asterisk represent adjusted odds ratio and the corresponding 95% confidence interval as well as statistically significant estimation with *p*-value < 0.05, respectively. Color of red(/blue) highlights the negative (/positive) effects of covarables on vaccine uptake. Ages 18-44y was as reference group for age variable, and primary or below was as reference group for education level variable in the regression analysis.

## Discussion

We explored changes in vaccine hesitancy and vaccine uptake over time for COVID-19 and predictors of intention to receive COVID-19 vaccine and decisions in Hong Kong from October 2021 through to December 2022, and in the midst of large wave of Omicron wave that a local Vaccine Pass scheme was implemented on 24 February 2022. Among adults ≥65 years of age vaccine hesitancy (any dose) dropped from 30% at the end of 2021 to 2.8% in mid-July 2022 after the large Omicron wave. On the other hand, in the same time period, hesitancy rates (any dose) changed from 18.7% to 3.1% for younger adults 18-44 years, while the rates similarly decreased from 17.8% to 2.8% for those 45-64 years. In the multivariable regression analysis, our findings suggested that the Vaccine Pass policy was positively associated with individual vaccination uptake during the time period from pre-announcement to full implementation stages of the policy. In addition to the Vaccine Pass policy, other statistically sigificant predictors of vaccine uptake included age group, underlying medical conditions, higher perceived susceptibility of contracting COVID-19, higher trust in government in controlling COVID-19 and higher education in our Chinese population.

We found a more substantial overall decrease in vaccine hesitancy in older adults (≥65y) compared to younger adults (18-44y and 45-64y) following the implementation of Vaccine Pass Policy in Febrary 2022. In fact, vaccination intention can be a function of the disease risk and the vaccine effectiveness as well as vaccine safety (12). When risk of infection and risk of severe disease are perceived to be low, because of successful control measures or for other reasons, it could be viewed as a rational decision to postpone vaccination to avoid any risk of vaccine-related adverse effects (12). This might somewhat explain vaccine hesitancy through to the end of 2021 in Hong Kong particularly in older adults with underlying chronic medical conditions who may have been overly concerned about the risk of adverse events following vaccination (13). Older adults’ vaccine hesitancy could have been further intensified by perceiving the effectiveness of containment measures which the government regularly reassured the public would continue to be successful before the Omicron outbreak occurred (13).

Vaccine passport schemes have been suggested to incentivize vaccination (14) so as to reducing individual vaccine hesitancy and increasing vaccine uptake (15, 16), despite mixed views of public regarding potential benefit (14) and harms in terms of social equality, social isolation and health disparities among those choosing to remain unvaccinated (17, 18). The coercion implicit in a vaccine passport scheme has also been discussed in terms of informed consent. One of the three principles of informed consent to receive a medical procedure is that it is freely and voluntarily given (19). We believe the expansion of the vaccine pass to require a third dose might be considered disproportionate for younger adults for whom the risk of severe COVID-19 is much lower (20), noting that a very high level of two-dose vaccine uptake (≥80%) was already achieved in younger adults by the end of 2021 well before the vaccine pass policy was announced (Figure 2) (21). For older people who might have more difficulties in deliberating the risk and benefit of vaccination and require more social support for vaccination decisions (13), a vaccine mandate or vaccine passport scheme could be considered to increase vaccination rates. Such personal experience with the vaccination can help to reduce unrealistic expectations about vaccine side effects. In this context, the vaccine scheme might have its merit. However, we believe that vaccination as a medical decision should be implemented on the basis of full informed consent, without coercion, in order to reduce psychological resistance and maintain long-term trust in science and health authorities especially for individuals who prefer autonomy to make their medical decisions.

Indeed, it may be important to consider the rationale for a two-dose vaccine passport somewhat separately from a three-dose vaccine passport. Many locations around the world, including Ireland (22), United Kingdom (23), United States (24) and South Korea (25) introduced versions of a two-dose passport, with most being relaxed by early 2022 once two-dose uptake had reached a high level. On the other hand, we have identified a few other locations which implemented a requirement for three doses in all adults (in Singapore, Israel) or in health care workers (in Australia). While a third dose undoubtely increases protection against severe COVID to an even higher level than than provided by two doses, the necessity for coercing receipt of the third dose is less clear in our opinion.

Nevertheless, the Vaccine Pass was not applicable to older adults in residential care homes. This might attribute to the relatively low vaccination coverage in care home residents when the Omicron variant began to spread locally, and this accounted for a large number of COVID-19 deaths (26). In fact, the vaccination uptake rate of care home residents was observed to be 12.3% in February 2022. With respect to the high COVID-19 mortality impact of Omicron BA.2 in early 2022, COVID-19 deaths in care home residents accounted for approximately 40% of the 11,000 COVID-19 deaths that occurred, and only about one in ten of these COVID-19 deaths among care home residents occurred in individuals who had completed two or more doses of COVID vaccination.

In our study, individuals reporting doctor-diagnosed chronic conditions had a lower odds of vaccine uptake. These associations were similarly observed in a recent study among older adults in mainland China, in which older people might perceive their chronic conditions as vaccine contraindication due to vaccine-safety scares coupled with deficiency in health literacy (27). Likewise, our findings were also consistent with a local study where a substantial proportion of the Hong Kong population had the misconception that individuals with chronic conditions should not be vaccinated (26). In fact, our study respondents who hesitated or refused to take their first dose had reported their major concerns about adverse reactions to vaccination, vaccine safety as well as poor health or chronic illness (data not shown). To reduce vaccine hesitancy at the community level, future vaccination programs may need more practical advice about what individuals can do to prepare for unanticipated vaccine-safety scares and to maximize uptake rates (28). On the other hand, we believe that increases in public trust regarding vaccine safety would be essential (29).

Our empirical findings confirmed the positive association between trust in government and vaccination uptake that was revealed in recent systematic reviews (30). This might also reflect the critical role of the government-public relationship in shaping vaccine acceptance and success of the vaccination programme. The inadequate trust in government-public relationship could result in increased vaccine hesitancy by concerning the reliability about the vaccine information sourced from the government (31). This also reminds us that trusted sources of information and guidance are fundamental to disease control (28). However, in a unique scenario for Hong Kong compared to other locations, health authorities and local health experts continued to reassure the public that COVID-19 could be contained successfully by non-pharmaceutical interventions (32). This may have affected the personal risk-benefit assessment of COVID-19 vaccination in some individual. In contrast, for example, health officials in Singapore made it clear by June 2021 that the successful containment of COVID-19 with non-pharmaceutical interventions was only temporary, and vaccination was essential for older adults in advance of the planned transition away from containment towards the end of 2021 (32, 33).

We also found that COVID-19 vaccine uptake was not homogenous across educational subgroups. We found that higher educational attainment was significantly associated with increased receipt of COVID-19 vaccination. This was similarly observed nearly in all countries and was most pronounced in high-income countries overall, based on the University of Maryland Social Data Science Center Global COVID-19 Trends and Impact Survey in 2022 (34). Continued inequality monitoring for population and key subgroups may help to inform targeted actions for more equitable uptake of vaccines (34).

Several limitations are acknowledged. First, our data were collected from cross-sectional telephone surveys which did not allow us to examine within-person psycho-behaviorial changes throughout the pandemic. The temporal associations of the psycho-behaviorial covariates (such as risk perception and trust in government) with vaccination uptake examined in our study might be susceptible to reverse causality (26, 35). For example, vaccine policy issued by the government might lead to mistrust and negative views toward government in the unvaccinated group (26). Second, there might be residual confounding of the effect of the Vaccine Pass policy due to other factors that we did not measure. The introduction of the Vaccine Pass policies occurred during and after a large omicron wave with high mortality (Figure 1), and thus it is particularly challenging to disentangle the true association for policy changes on vaccination uptake under those rapid contextual changes in COVID-19 infections and mortality that could affect vaccination decisions.

In conclusion, we found that vaccine uptake appeared to have increased by a greater extent among older adults during the timing of post-announcement and full implementation of the Vaccine Pass policy, under the contextual changes in the midst of large wave of Omicron wave with high mortality in Hong Kong in 2022. While the third dose undoubtedly increases protection against severe COVID-19, the necessity for inclusion of a third dose requirement in the Vaccine Pass policy merits further discussion. Since the withdrawal of the Vaccine Pass policy in late 2022 the uptake of further boosters has been sluggish, and improving voluntary booster uptake in older adults in future years should be a public health priority.

## Contributors

The study was designed by IW and BJC. The survey tools were developed by JX, BJC and QL. Funding was obtained by MYN, BJC and QL. Data analyses were done by IW, CW, NM and AD. IW and BJC wrote the first draft of the manuscript. All authors interpreted data, provided critical review and revision of the text and approved the submitted manuscripts.

## Data Availability

Data will be made publicly available after publication of peer-reviewed article

## Acknowledgements

We thank Julie Au for administrative support. We received funding from the Health Bureau, The Government of the Hong Kong Special Administrative Region (Ref. No.: COVID19F04; COVID19F11), and by the Theme-Based Research Scheme T11-705/21-N of the Research Grants Council of the Hong Kong Special Administrative Region, China. B. J. C. is supported by a RGC Senior Research Fellow Scheme grant (HKU SRFS2021-7S03) from the Research Grants Council of the Hong Kong Special Administrative Region, China.

