## Supplementary Materials for "Assessment of the impact of the Vaccine Pass Policy on COVID-19 vaccine hesitancy and uptake among Chinese adults in Hong Kong"

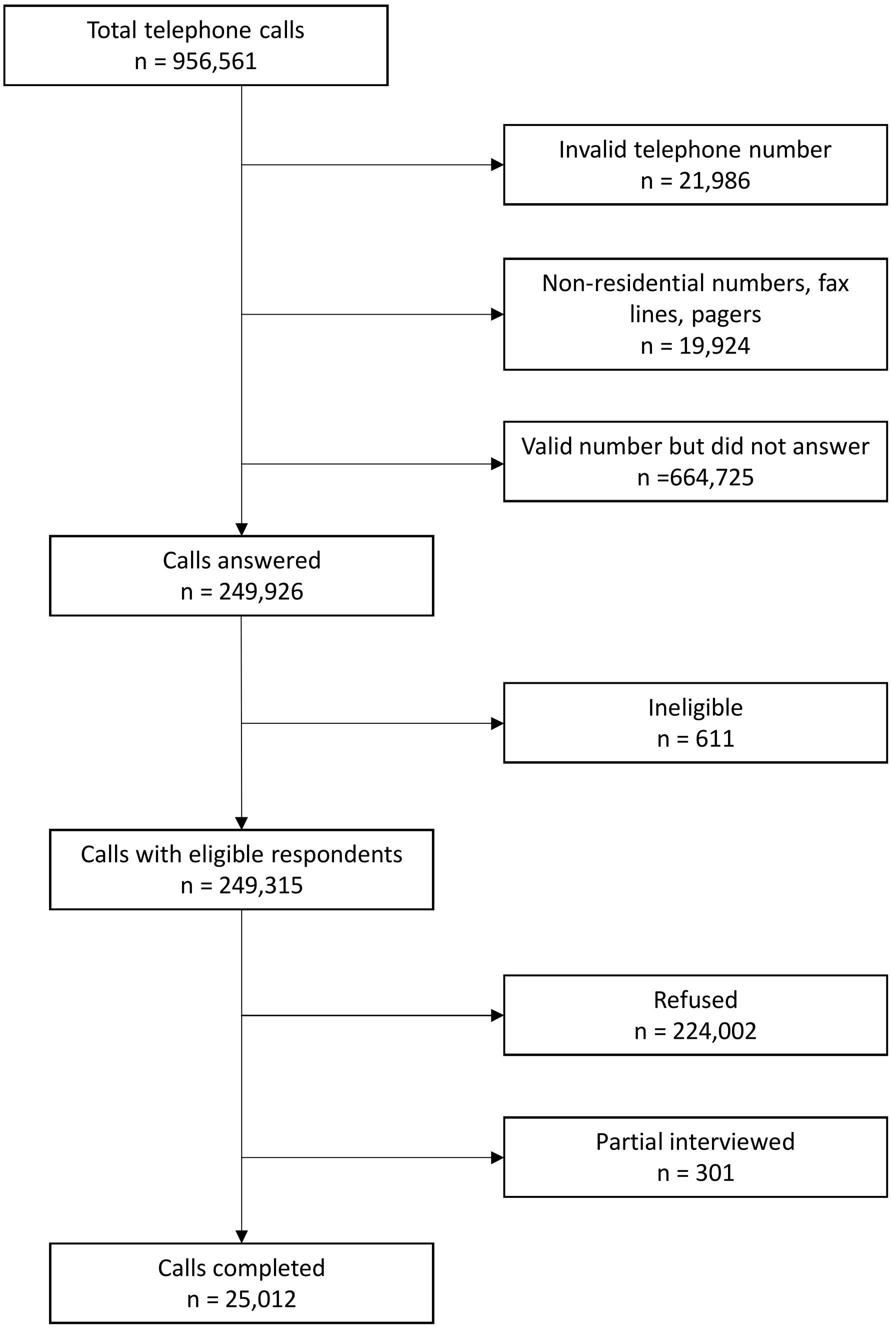


**Supplementary Figure 1.** Flow chart of Participant recruitment from October 2021 to December 2022

**Supplementary Table 1.** The Hong Kong Vaccine Pass scheme

| **Date in 2022** | **Key events related to Vaccine Pass** | **Descriptions, if any** |
|---|---|---|
| Feb 8 | Announcement: Vaccine Pass plan | |
| Feb 21 | Announcement: Vaccine Pass policy details | All HK residents shall show proof that they have received received at least one dose of COVID-19 vaccination to enter government venues and 24 other types of private premises including all catering business premises (including bars or pubs), amusement game centres, bathhouses, fitness centres, places of amusement, places of public entertainment, party rooms, beauty parlours and massage establishments, clubs or nightclubs, karaoke establishments, mahjong-tin kau premises, club houses, sports premises, swimming pools, cruise ships, event premises, religious premises, barber shops or hair salons, shopping malls, department stores, supermarkets, markets, and hotels or guesthouses (only applicable to staff members). Individuals are required to show their vaccine pass for digital scanning and record. Vaccine Pass is a QR code on a paper vaccination certificate or the LeaveHomeSafe app that contains COVID-19 vaccination history. Individuals who have not received at least one dose are prohibited from entering government buildings and the other 24 types of designated premises listed above, or otherwise may face a maximum fine of HK$10,000 (US$1280). Under the arrangement of the Vaccine Pass, a person who is medically unsuitable for vaccination is required to obtain an Exemption Certificate issued by a doctor.* |
| Feb 24 | Implementation: **Phase 1** Vaccine Pass | By April 29, individuals aged 12 and over are required to have received at least one vaccine dose. |

| | | |
|---|---|---|
| Apr 30 | Implementation: **Phase 2** Vaccine Pass | During the period, individuals aged 18y and over are required to have received at least two doses. Individuals aged 12-17y are required to have received their 2nd dose when their 1st dose was received more than 6 months earlier unless they can show proof of a laboratory-confirmed infection within the last 6 months. |
| May 21 | Announcement: expand Vaccine Pass coverage to designated public-sector healthcare premises including specialist outpatient clinics and public dental clinics but excluding emergency services | |
| May 31 | Implementation: **Phase 3** Vaccine Pass | By May 30, individuals aged 12y and over are required to have received at least two doses. Those individuals aged 12y are required to have received their 3rd dose when their 2nd dose was received more than 6 months earlier unless they can show proof of a laboratory-confirmed infection within the last 6 months. |
| Jun 13 | Implementation: expand Vaccine Pass coverage to designated public-sector healthcare premises | |
| Sep 30 | Implementation: **Stage 1** Vaccine Pass for children aged 5-11y | All children aged 5 to 11y are required to have received at least one dose |
| Nov 30 | Implementation: **Stage 2** Vaccine Pass for children aged 5-11y | All children aged 5 to 11y are required to have received at least two doses |
| Dec 29 | Vaccine Pass scheme ceased | |

*Of note, approximately 40,000 exemption certificates were issued (for reference, the population of Hong Kong is 7.3 million). On 27 September 2022 the government invalidated more than 20,000 exemption certificates issued by seven doctors, and banned those seven doctors from issuing any further exemption certificates. Several doctors and more than 20 patients were arrested in connection with allegedly fraudulent certificates, and at least two of the seven doctors subsequently faced criminal charges in court.
